# Preoperative serum C-reactive protein and cholinesterase levels as risk factors of difficult laparoscopic cholecystectomy and severity of acute calculous cholecystitis: a retrospective observational study

**DOI:** 10.64898/2026.02.25.26347116

**Authors:** Cai-Qin Kang, Liang-Ping Chen, Yu-Xian Wang

## Abstract

**Background:** Early laparoscopic cholecystectomy (ELC) is the standard treatment for acute calculous cholecystitis (ACC), but difficult laparoscopic cholecystectomy (DLC) remains a challenge. Predicting DLC and ACC severity is crucial for clinical decision-making.

**Methods:** This retrospective single-center study included 198 ACC patients who underwent ELC. Preoperative clinical, laboratory, and imaging data were analyzed. DLC was defined by operative time >90 min, conversion, or subtotal cholecystectomy. ACC severity was graded using TG18. Multivariate logistic regression identified independent predictors.

**Results:** DLC occurred in 81 (40.9%) patients; 102 (51.5%) had severe ACC. Serum cholinesterase (ChE) and CRP were independent predictors of DLC. CRP and male sex independently predicted ACC severity. Other markers (e.g., NLR, PCT) were not independently associated.

**Conclusion:** Preoperative ChE and CRP levels are reliable predictors of DLC, while CRP and male sex predict ACC severity. These findings support their use in risk stratification and surgical planning.

## Introduction

Cholelithiasis affects 10-15% of the general population, and acute calculous cholecytitis (ACC) is the most common complication [1]. According to the World Society of Emergency Surgery (WSES), ACC is the second cause of complicated intra-abdominal infections, accounting for 18.5% of the total number of cases [2]. Possible complications of ACC include perforation of the gallbladder, peri-gallbladder abscess, internal biliary fistula, biliary peritonitis, and acute cholangitis [3]. Early laparoscopic cholecystectomy (ELC) is becoming the gold standard of treatment in acute cholecystitis, since it offers better safety, shorter hospital stay, fewer perioperative complications, and lower financial burden [4, 5]. However, performing ELC in ACC may be challenging due to inadequate visualization of anatomical landmarks and difficulties of achieving a critical view of safety. It is reported that 36.7% of ELCs performed in emergency had a non-standard outcome, which are sometimes defined as difficult laparoscopic cholecystectomy (DLC), including conversion to open surgery, subtotal cholecystectomy, bile leak, and prolonged postoperative hospital stay. Most of these cases were related to the severity of inflammation of the gallbladder, and difficult dissection of the Calot Triangle [6–8]. According to Tokyo guidelines 2018 (TG18), ELC should be performed based on ACC severity grading, and be carefully used in special clinical settings [9]. Preoperative identification of the severe cases of ACC is important to clinical decision-making for cholecystectomy.

In recent years, there has been considerable research on biomarkers, methods, and models for predicting the severity of acute cholecystitis. The TG18 guidelines proposed a method for grading the severity of the condition using clinical assessment, white blood cell count (WBC), C-reactive protein (CRP), and other clinical data [9]. However, a systematic meta-analysis has found that this grading system does not effectively predict patients’ surgical risks before an ELC [10]. Many biomarkers, such as CRP [11, 12], procalcitonin (PCT) [13], neutrophil-to-lymphocyte ratio (NLR), platelet-to-lymphocyte ratio (PLR), systemic inflammatory index (SII), and other laboratory markers [14], in combination with imaging results from ultrasound and CT have been studied as preoperative predictive indicators [15–17]. More recently, artificial intelligence was used for classification of cholecystitis severity [18, 19]. Those studies have shown that these indicators—either individually or in combination—can provide certain value for preoperative assessment of ACC severity or DLC. However, the conclusions of the studies are inconsistent, and more clinical evidence is needed to identify appropriate indicators and models that can be used in therapeutic management.

Cholinesterase (ChE) is synthesized mainly in hepatocytes, and is involved in the regulation of the balance between the proliferation and death [20, 21]. Recent studies have demonstrated that serum levels of ChE is related to pesticide exposure, inflammation, injury, infections, and malnutrition [22]. The serum ChE level is measured routinely in many clinical laboratories, and is a potential predictor for clinical comes of various diseases, such as inflammatory diseases, cancers, and cardiovascular diseases [23–25]. However, whether serum levels of ChE are associated with the severity of ACC, or the outcomes of ELC, is unclear.

In the present study, we aimed to analyze the value of preoperative factors, including the serum ChE level, in predicting difficult laparoscopic cholecystectomy and severity of acute calculous cholecystitis.

## Materials and methods

This is a single center, retrospective observational study. Patients visited at Dingxi People’s Hospital (Gansu, China) for acute cholecystitis and underwent cholecystectomy between July 2023 and June 2025 were included. All patients were admitted in emergency care, and aged over 18 years. Data were collected from electronic medical records and operatory protocols. Laboratory parameters were recorded at the first day of hospitalization admission. Ultrasonography and/or computed tomography (CT) were performed before the operation, and used to document the presence of calculi, the thickness of the gallbladder walls, and common biliary duct diameter. Pathologic examines were performed after the operation, and two experienced pathologists carefully reviewed the results independently. Comorbidities, time elapsed from the onset of symptoms to presentation, and clinical signs were also assessed at admission. Patients with a concomitant diagnosis of pancreatitis, obstructive jaundice, ascending cholangitis, tumors, hematological malignancies, or autoimmune diseases were excluded.

Difficult laparoscopic cholecystectomy was defined according to the presence of any of the following factors: operative time more than 90 min, drain placement, bile or stone spillage, conversion to open surgery, or subtotal cholecystectomy. Severity of ACC were classified according to pathological results and graded by the TG18 [9].

Continuous variables are presented as medians with ranges or means with standard deviation. For categorical variables, percentages of patients in each category were analyzed. Comparisons between groups were performed using the Mann-Whitney test, chi-square test, or two-sample *t*-test, depending on variable characteristics. Univariate analysis and multivariate logistic regression were performed to determine whether the variables were independent factors associated with DLC or severity of ACC. All analyses were performed using SPSS, version 26.0 (IBM, Armonk, NY, USA). Tests were two-sided, with significance assessed at the 0.05 level.

The ethics committee of Dingxi People’s Hospital approved this retrospective cohort study. Written informed consent was waived owing to the retrospective nature of the data retrieval.

## Results

### Study Cohort

The study retrospectively included 198 patients with ACC. Eighty-four patients (42.4%) were men, and 114 (57.6%) were women. The median age was 53 years (range, 19-83 years). A total of 51 patients (25.8%) had diabetes mellitus or other complications. Among all the patients, there were 81 (40.9%) DLC cases and 102 (51.5%) severe cases. No conversion to open surgery was identified among the patients. Compared to non-DLC patients, DLC patients had significantly higher levels of alkaline phosphatase (ALP), total bilirubin (TBil), direct bilirubin (DBil), total bile acids (TBA), CRP, PCT, and hospital stay time, but significantly lower levels of ChE. Compared to non-severe patients, severe patients had significantly longer duration of symptoms, more times of pain in 24 hours, higher levels of of white blood cell (WBC) count, monocytes count, neutrophils count, monocyte-to-lymphocyte ratio (MLR), neutrophil-to-lymphocyte ratio (NLR), alanine aminotransferase (ALT), alkaline phosphatase (ALP), direct bilirubin (DBil), total bile acids (TBA), CRP, more patients with thickness of bladder walls > 3mm, and longer hospotal stay time, but significantly lower levels of CH (Table 1). Levels of ChE in different groups were inllustrated in Fig 1.

**Fig 1.**
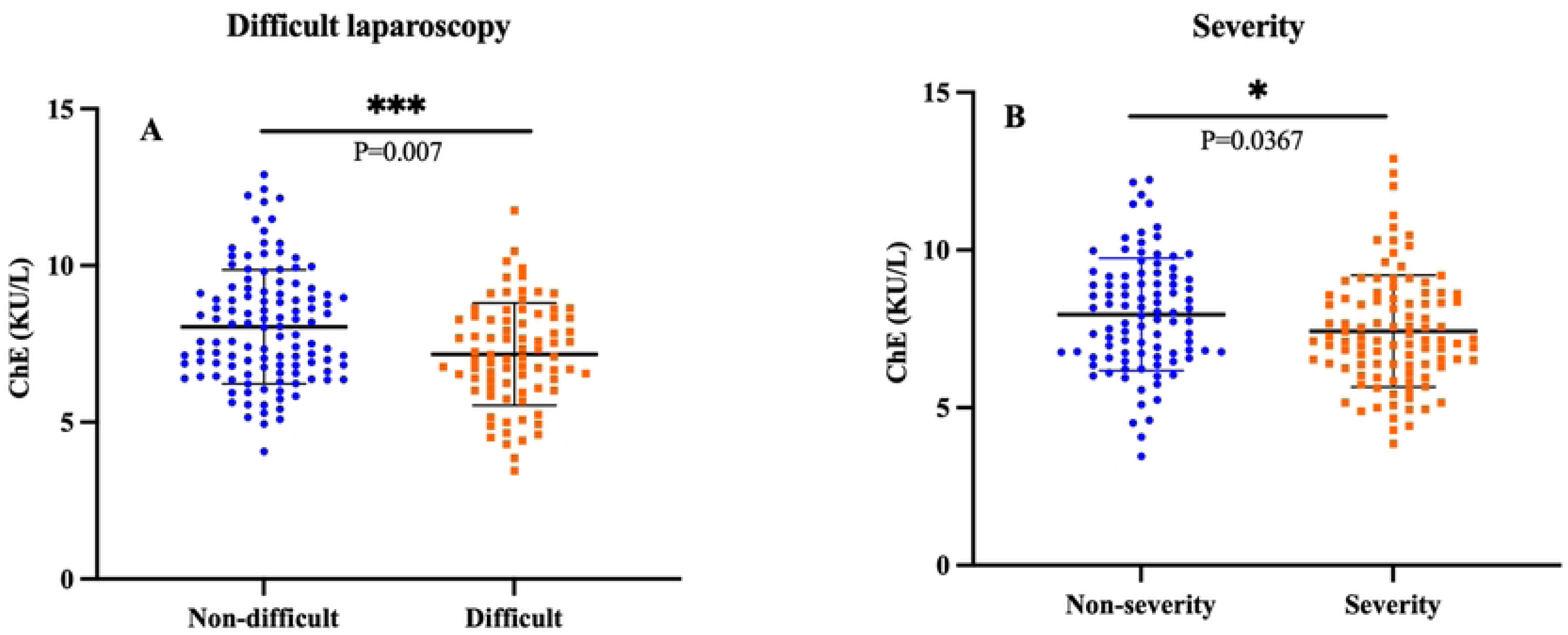
Serum Cholinesterase (ChE) levels among different groups. A: ChE levels in non-difficult and difficult laparoscopy groups showed that ChE levels were significantly decreased in the difficult group *(P* = 0.007). B: ChE levels in non-severity and severity groups showed that ChE levels were significantly decreased in the severity group (*P* = 0.0367). The middle lines in the figure represent the median values; the upper and lower lines represent the 25^th^ and 75^th^ percentile.

**Table 1.**
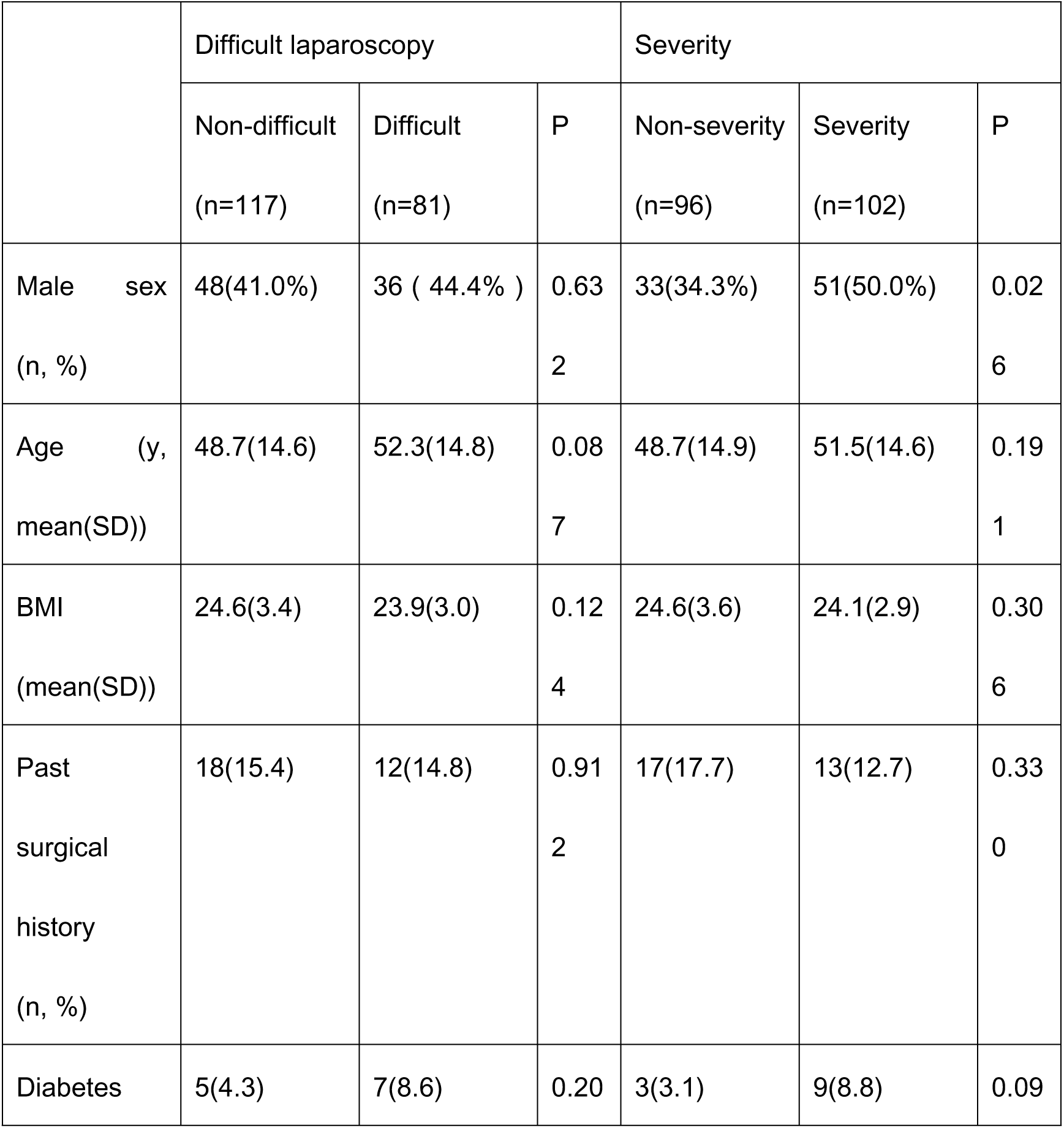
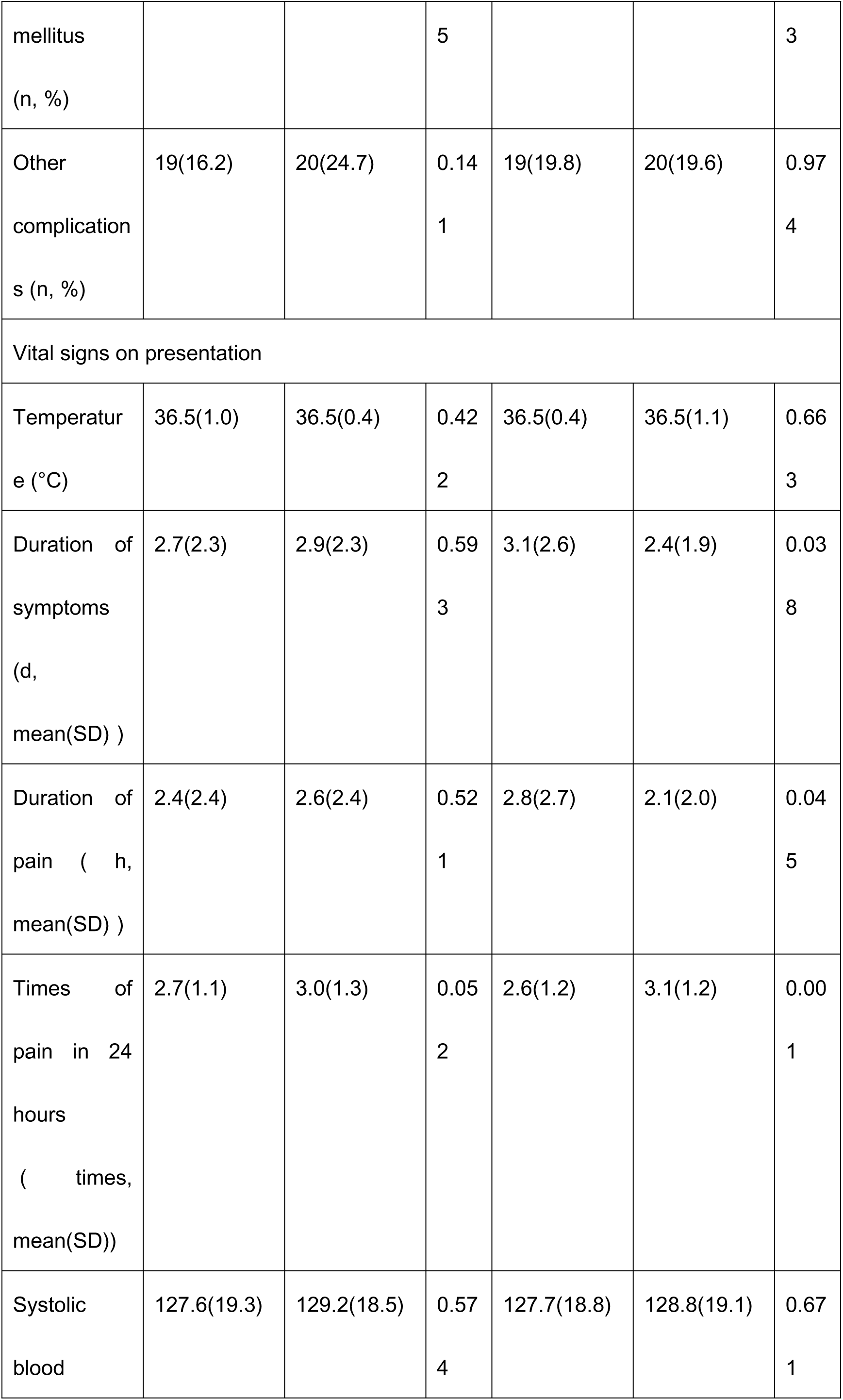
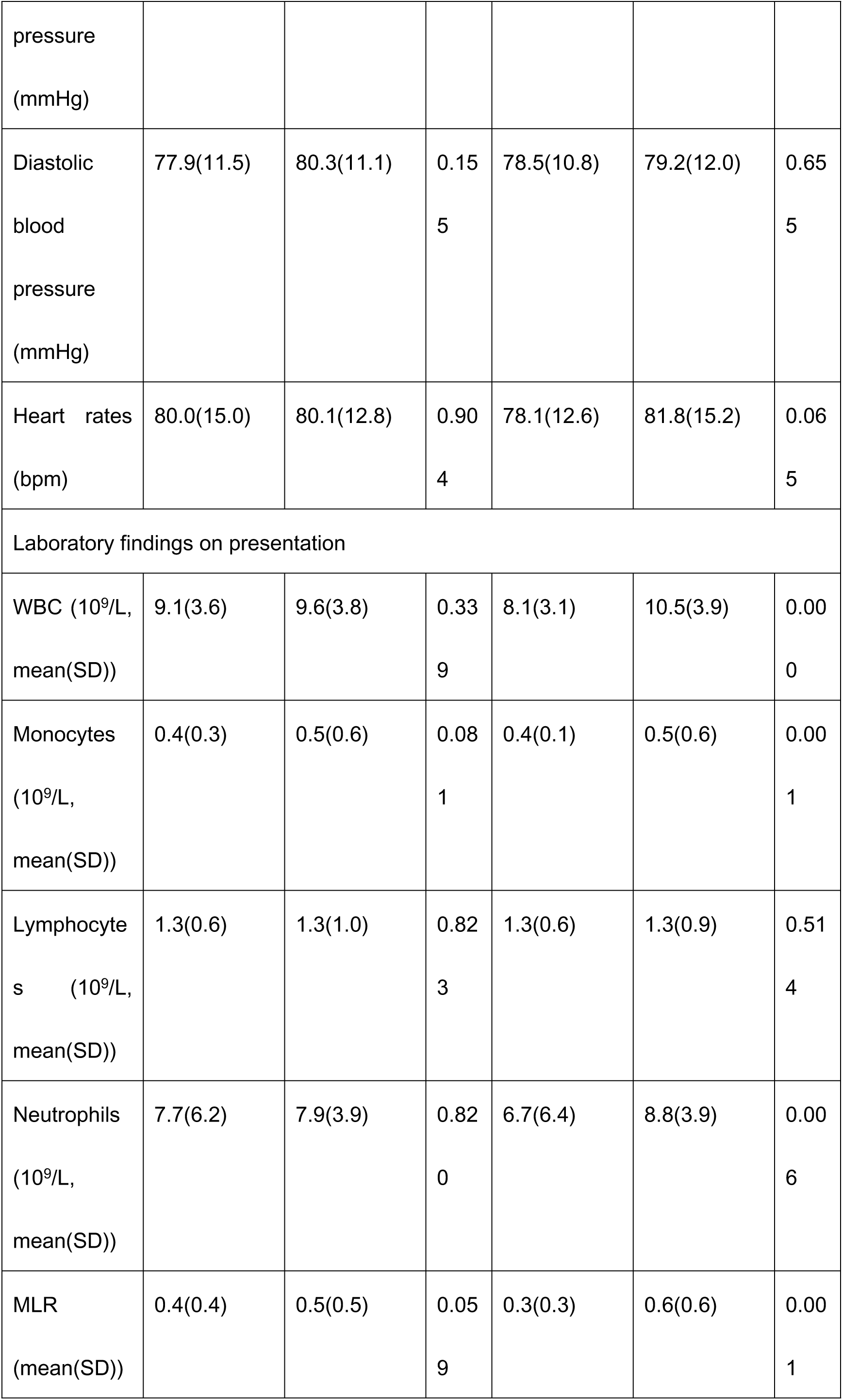
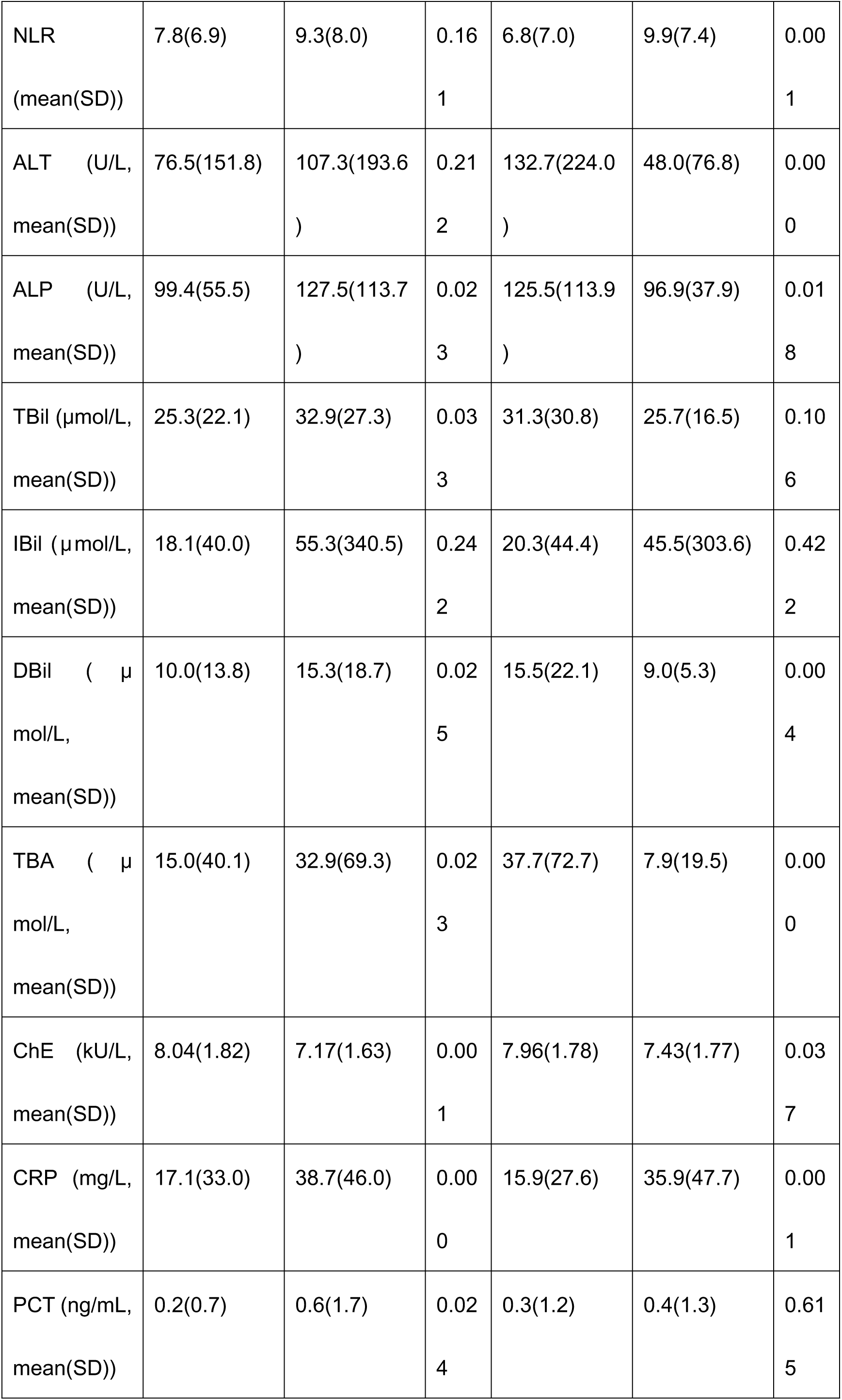
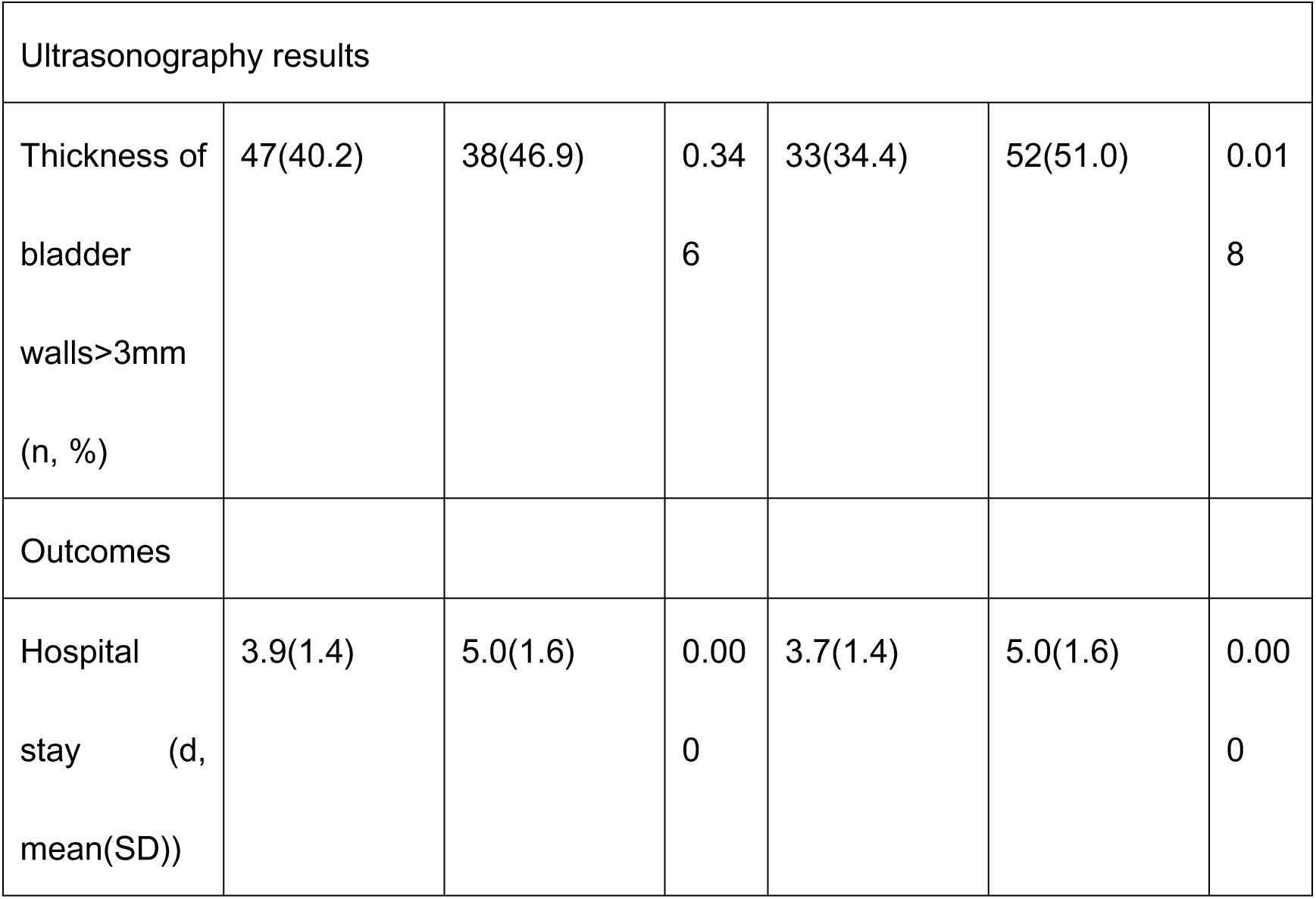
Baseline demographic and clinical characteristics.

### Univariate analysis

In the univariable Logistic regression analysis, higher levels of ALP (odds ratio [OR], 1.004, 95% CI, 1.000-1.008, *P* = 0.036), TBil (OR, 1.013, 95% CI, 1.000-1.026, *P* = 0.043), DBil (OR, 1.022, 95% CI, 1.00-1.043, *P* = 0.040), and CRP (OR, 1.014, 95% CI, 1.006-1.023, *P* = 0.001) were each positively associated with DLC risk, but high levels of ChE was negatively associated with DLC risk (OR, 0.745, 95% CI, 0.624-0.888, *P* = 0.001). Other clinical and biochemical variables, including WBC, NLR, and PCT, did not exhibit statistically significant associations with DLC. For severity of ACC, male sex (OR, 1.909, 95% CI, 1.077-3.385, *P* = 0.027), more times of pain in 24 hours (OR, 1.498, 95% CI, 1.162-1.931, *P* = 0.002), WBC (OR, 1.224, 95% CI, 1.117-1.342, *P* = 0.000), monocytes count (OR, 1.909, 95% CI, 1.077-3.385, *P* = 0.027), neutrophils count (OR, 1.118, 95% CI, 1.035-1.208, *P* = 0.005), MLR (OR, 4.906, 95% CI, 1.726-13.948, *P* = 0.003), NLR (OR, 1.067, 95% CI, 1.021-1.115, *P* = 0.004), CRP (OR, 1.015, 95% CI, 1.005-1.024, *P* = 0.002) and thickness of bladder wall > 3 mm (OR, 1.985, 95% CI, 1.120-3.521, *P* = 0.019) were each positively associated with severity of ACC. However, duration of symptoms (OR, 0.876, 95% CI, 0.772-0.994, *P* = 0.040), duration of pain (OR, 0.884, 95% CI, 0.783-0.998, *P* = 0.047), levels of ALT (OR, 0.995, 95% CI, 0.992-0.998, *P* = 0.003), ALP (OR, 0.995, 95% CI, 0.991-0.999, *P* = 0.029), DBil (OR, 0.965, 95% CI, 0.939-0.993, *P* = 0.013), TBA (OR, 0.983, 95% CI, 0.972-0.994, *P* = 0.003), and ChE (OR, 0.844, 95% CI, 0.718-0.991, *P* = 0.039) were negatively associated with severity of ACC. Other variables did not statistically associated with severity of ACC. Varables with *P* value bellow 0.100 were summarized in Table 2.

**Table 2.**
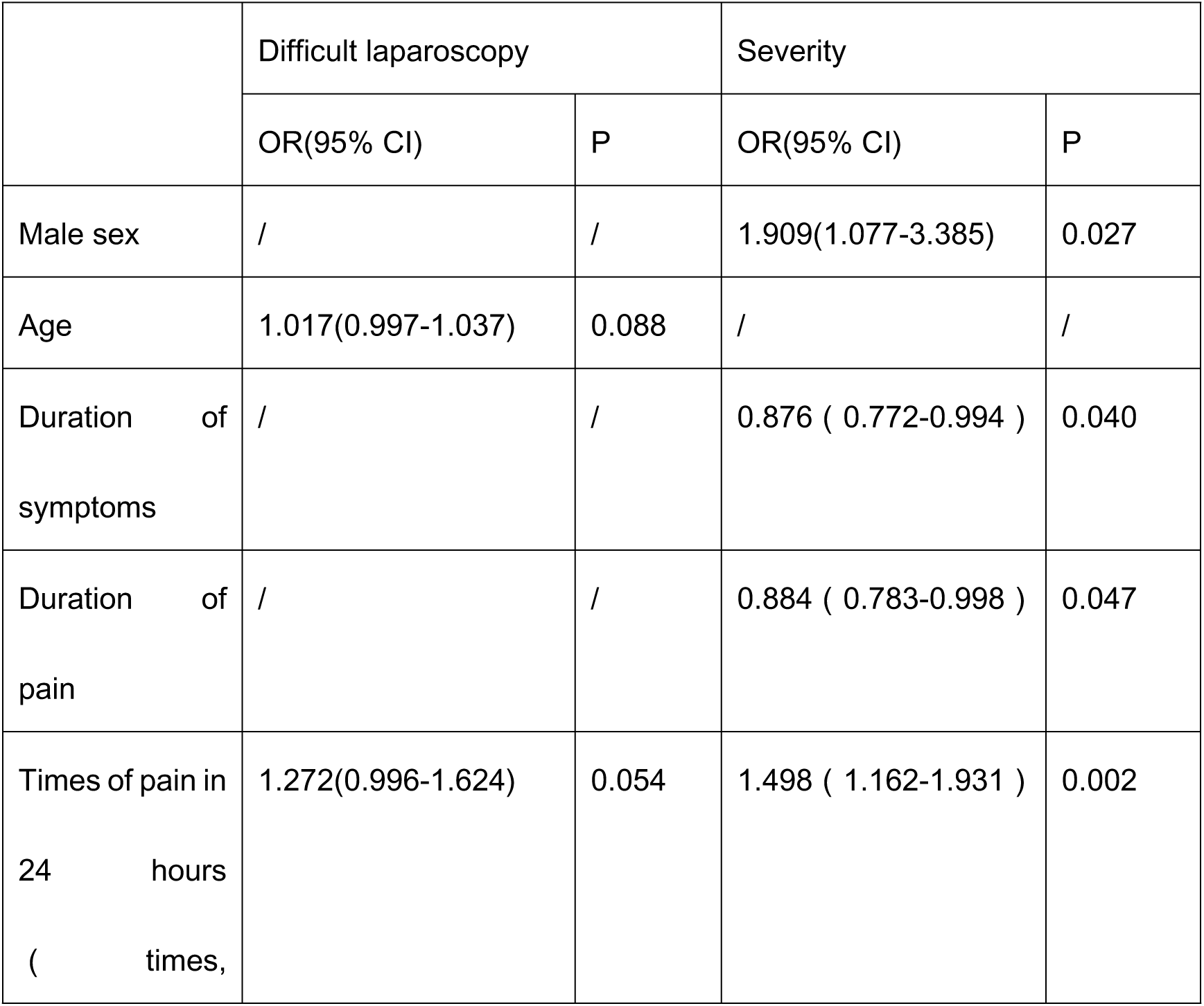
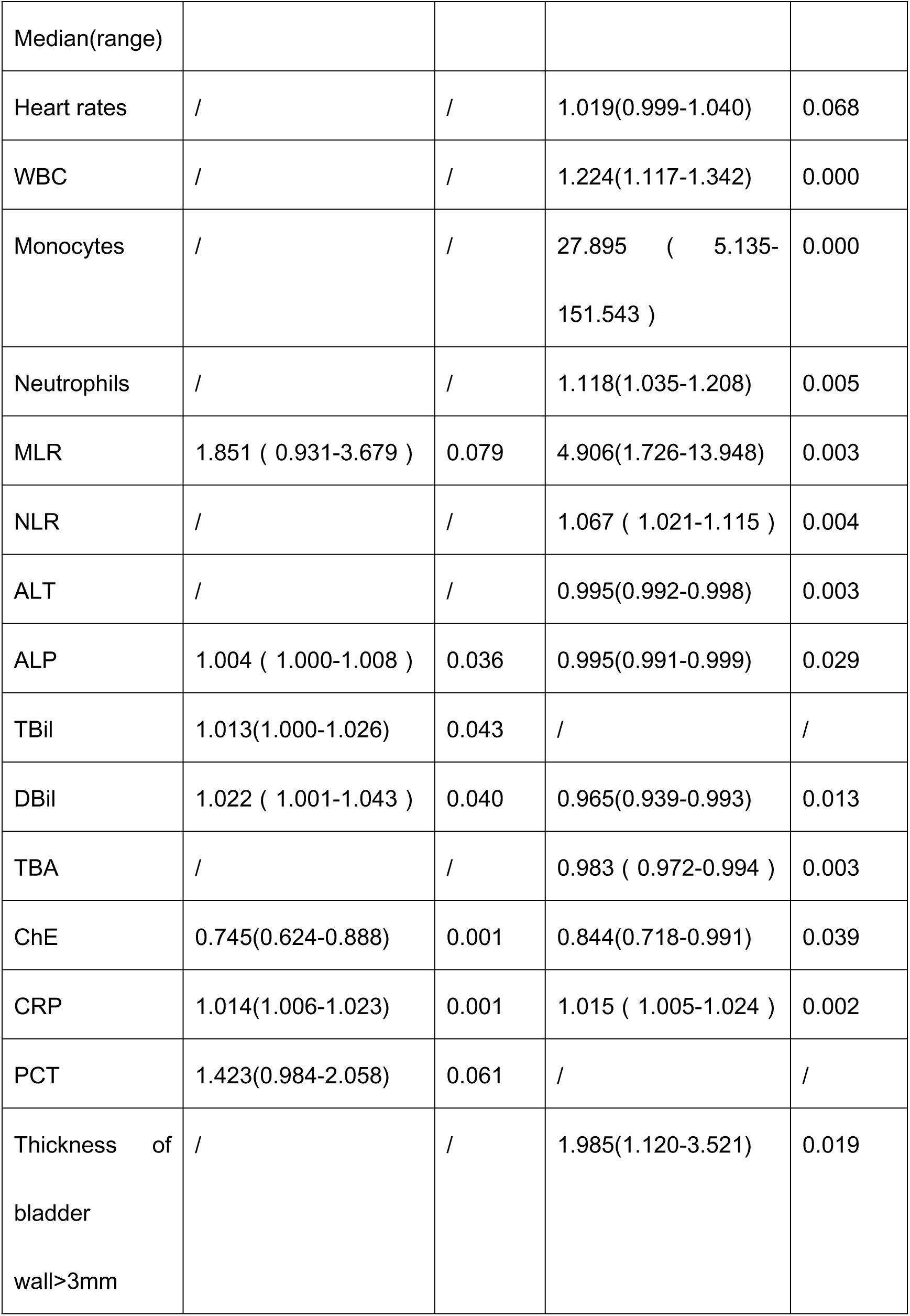
Univariant logistic regression analysis.

### Multivariate analysis

Factors associated with increased odds of DLC included ALP, TBil, DBil, ChE, and CRP. In the multivariate analysis, only ChE (OR, 0.819, 95% CI, 0.679-0.987, *P* = 0.036) and CRP (OR, 1.011, 95% CI, 1.003-1.020, *P* = 0.010) were independently associated with DLC. CRP is a risk factor and ChE is a protective factor.

Factors associated with increased odds of severity included male sex, duration of symptoms, duration of pain, times of pain in 24 hours, WBC, monocyte count, neutrophil count, MLR, NLR, ALT, ALP, DBil, TBA, CRP, and the thickness of bladder wall. However, in the multivariate analysis, only male sex (OR, 2.132, 95% CI, 1.028-4.420, *P* = 0.042) and CRP (OR, 1.017, 95% CI, 1.002-1.031, *P* = 0.021) were independently associated with severity of ACC. The results were summarized in Table 3.

**Table 3.** Multivariant logistic regression analysis.

|  | Difficult laparoscopy |  | Severity |  |
| --- | --- | --- | --- | --- |
|  | OR(95% CI) | P | OR(95% CI) | P |
| Male sex | / | / | 2.132(1.028-4.420) | 0.042 |
| ChE | 0.819(0.679-0.987) | 0.036 | / | / |
| CRP | 1.011(1.003-1.020) | 0.010 | 1.017(1.002-1.031) | 0.021 |

## Discussion

ACC is a common and frequent condition, accounting for a significant proportion of emergency surgical patients [9]. Laparoscopic cholecystectomy (LC) is the most common treatment for ACC. Although LC is safe and effective, a certain percentage of patients develop difficult laparoscopic cholecystectomy (DLC), and the severity of ACC directly influences clinicians’ therapeutic decisions. Accurately predicting both the severity of ACC and the likelihood of DLC is therefore of great clinical value: it enables early prevention of adverse outcomes, shortens hospital stays, improves treatment efficacy, and reduces overall costs.

Although several preoperative predictors of difficult laparoscopic cholecystectomy (DLC) have been reported, their findings remain inconsistent. Alburfakan et al. found that the Charlson Comorbidity Index (CCI) is a reliable predictor of DLC [26]. Gupta et al. developed a scoring system based on history of hospitalization, palpable gall bladder, impacted stone and gall bladder wall thickness, with positive predictive values of 90% and 88% for easy and difficult cases respectively [27].

In addition to clinical features and imaging findings, laboratory parameters, especially inflammatory markers and liver-function tests, are also evaluated for predicting DLC. WBC, CRP, PCT and NLR have all been reported to predict both DLC and disease severity [28–31]. In our study, ALP, TBil, DBil, TBA, ChE, CRP and PCT differed significantly between DLC and non-DLC patients (*P* < 0.05). However, multivariate logistic regression identified only ChE and CRP as independent predictors of DLC. Our findings support the predictive value of CRP for DLC, but do not support a predictive role for other markers.

Studies of predictors for ACC severity have shown that the extent of disease is closely linked to both systemic and local inflammation. TG18 grades acute cholecystitis into three levels, basing the stratification largely on the degree of inflammation and the presence of organ involvement [9]. Because a large number of inflammatory biomarkers exist, published findings vary. Nevertheless, an increasing body of evidence supports CRP as a key predictor of severity. In the present cohort no patient met Grade III criteria, so we defined Grade II cases as “severe” and all others as “mild.” The two groups differed significantly in multiple clinical and laboratory variables (*P* < 0.05). Univariate analysis identified the following factors as associated with severity: male sex, symptom duration, pain duration, number of pain episodes within 24 h, WBC, monocyte count, neutrophil count, MLR, NLR, ALT, ALP, DBil, TBA, CRP, and gall-bladder-wall thickness. However, multivariate logistic regression revealed that only male sex (OR = 2.132, *P* = 0.042) and CRP (OR = 1.017, *P* = 0.021) were independently related to ACC severity—findings consistent with TG18 and previous reports. Although NLR and PCT are widely recognized systemic inflammatory markers and some earlier studies have suggested they predict ACC severity, our data do not support their independent predictive value.

Cholinesterase (ChE) is an α-glycoprotein synthesized by the liver and rapidly released into the bloodstream, and has been extensively studied across a wide range of clinical settings. Elevated ChE levels are associated with fatty liver disease, obesity, and metabolic syndrome [32], whereas reduced levels are characteristically observed in acute or chronic hepatic injury, liver cirrhosis, tumor metastasis to the liver, malnutrition, and systemic inflammation [33–35]. A low serum ChE concentration may mirror an underlying state of systemic inflammation—an important hallmark of severe acute cholecystitis (ACC). Pro-inflammatory cytokines such as interleukin-6 (IL-6) and tumour necrosis factor-α (TNF-α) are known to suppress hepatic ChE synthesis; this inflammatory milieu therefore compromises the liver’s capacity to produce ChE and drives circulating levels downward [23]. In addition, decreased ChE frequently reflects malnutrition, which itself impairs hepatocyte function and restricts synthesis of export proteins including ChE. Low ChE may also indicate primary hepatic dysfunction in patients with cholecystitis. Together these factors shape the clinical presentation of ACC, increase the technical difficulty of laparoscopic cholecystectomy (LC), and ultimately raise the likelihood of a difficult laparoscopic cholecystectomy (DLC).

This study has several limitations. It is a single-center retrospective analysis. The variables examined were restricted to data available at the time of admission in the electronic medical record. No patients with Grade III severity were included, and optimal cut-off values for the investigated biomarkers in ACC have not been firmly established. Nevertheless, our findings further underscore the importance of CRP in predicting both DLC and ACC severity, and highlight the potential value of ChE as a predictor of DLC, thereby providing useful insights for future investigations.

In conclusion, this study demonstrates that CRP and ChE are independent predictors of DLC, while CRP and male sex are independent predictors of ACC severity. To enhance the prediction of both DLC and ACC severity, multicenter prospective studies incorporating artificial intelligence are likely to provide more valuable information and more specific guidance for clinical practice.

## Data Availability

All relevant data are within the manuscript and its Supporting Information files.

